# Clinically significant estimated glomerular filtration rate decline is common in patients receiving immune checkpoint inhibitors: implications for long-term cancer survivors

**DOI:** 10.1101/2020.12.18.20248471

**Authors:** Donald F. Chute, Sophia Zhao, Ian A. Strohbehn, Nifasha Rusimbayala, Harish Seethapathy, Meghan Lee, Leyre Zubiri, Shruti Gupta, David E. Leaf, Osama Rahma, Zsofia D. Drobni, Tomas G. Neilan, Kerry L. Reynolds, Meghan E. Sise

## Abstract

**Purpose:** Immune checkpoint inhibitors (ICIs) are now indicated in more than one in three cancer patients. Acute kidney injury has emerged as an important complication of ICIs; however, there are no studies characterizing the long-term effects of ICIs on kidney function.

**Methods:** Retrospective cohort study of consecutive adult cancer patients treated with ICIs between 2010–2018 at two major cancer centers. The primary aim was to determine the composite outcome of incident or progressive CKD, defined as estimated glomerular filtration rate (eGFR) <60 mL/min/1.73m^2^ or a ≥30% decline in eGFR sustained >90 days among patients surviving ≥1 year. The secondary objective was to determine the proportion of patients experiencing rapid eGFR decline defined as >3mL/min/1.73m^2^ per year decline.

**Results:** A total of 5004 adult patients with cancer (mean [SD] age, 64 [13] years; 2699 [54%] male; 4529 [91%] White non-Hispanic) were included, and 2563 surviving ≥1 year were analyzed. During a median follow-up of 688 days (interquartile range, 496 to 1031), the overall event rate of the primary composite outcome was 17% and 20% at 3 and 5 years, respectively. Fine-Gray subdistribution multivariable hazard mode demonstrated that age and coronary artery disease were each independently associated with the primary outcome, adjusted HR (95% CI) 1.12 (1.06, 1.19) per 5 years and 1.27 (1.00-1.62), p < 0.001 and p=0.49, respectively.

**Conclusion:** New onset CKD or 30% eGFR decline is common in patients receiving ICIs who survive ≥1 year. As ICIs are increasingly used in earlier stage cancers, the impact of long term kidney function decline among survivors is extremely important. Additional studies are needed to determine the risk factors and potential modifiers for eGFR decline among patients receiving ICIs.

## Introduction

Initially approved in 2010 for patients with metastatic melanoma, immune checkpoint inhibitors (ICIs) are now approved for more than 17 different cancer types.^1^ Enhancing anti-tumor T-cell immunity with ICIs such as anti-programmed death 1 (PD-1), anti-programmed death ligand 1 (PDL1), anti-cytotoxic T-lymphocyte-associated protein 4 (CTLA-4) can lead to durable responses and even cures in subsets of with metastatic cancers. However, although ICIs augment the immune system’s ability to eradicate neoplastic cells, they can also lead to immune-related adverse events (irAEs), affecting as many as 60-85% of patients.^2, 3^ Renal irAEs affect 2-3% of patients treated with ICIs, typically causing acute tubulointerstitial nephritis, though autoimmune glomerular diseases have also been reported.^4-8^ ICI-associated nephritis typically occurs 2-6 months after starting therapy.^9^ Despite multiple studies examining the effects of ICIs on the kidneys acutely, few have examined their long-term effects on kidney function.

Currently more than one in three patients with cancer are eligible to receive ICIs and ongoing clinical trials are expanding use of ICIs for adjuvant therapy in early stage cancers, thus, the number of long-term survivors exposed to ICIs is expected to rise considerably.^1, 10-13^ Therefore, the study of medium to long term changes in health among surviving patients is extremely important. Several studies have established that chronic kidney disease (CKD) is independently associated with both overall and cancer-specific mortality among patients with cancer.^14, 15^ The aim of this study was to determine the rate of incident and progressive CKD in patients treated with ICIs who survived for at least 1 year.

## Methods

### Study design and Data Collection

We conducted a retrospective cohort study including all adults who received ICIs at Mass General Hospital and Dana Farber Cancer Institute between 2010-2018. The follow-up period began on the date of each patients’ first ICI exposure and continued until December 31, 2019. Cancer type and ICI start date were obtained from oncology infusion records. Covariates specifically included age, sex, race and ethnicity, cancer type, comorbidities, medications, prior cancer therapies, and laboratory test results were collected using the Research Patient Data Registry, the central data warehouse for Mass General Brigham healthcare. Receiving nephrotoxic cancer therapy was defined as at least one dose within 1 year prior to ICI start date (**Supplemental Table 1**). Baseline creatinine was an average of all creatinine values obtained within 6 months prior to ICI start. eGFR was calculated using the CKD-EPI equation.^16, 17^ Baseline comorbidities were defined using ICD-9/ICD10 codes. Death was determined by chart review or was assigned 45 days after the last laboratory datapoint in patients without laboratory studies for >6 months.

**Table 1.** Baseline characteristics. Baseline comorbidities and medications were defined at the time of ICI initiation. Baseline comorbidities were defined using ICD-9/ICD10 codes. Nephrotoxic chemotherapies had been prescribed within 1 year prior to ICI start. Blood cancers included myeloma, lymphoma and leukemia. Abbreviations: SD = standard deviation, eGFR = estimated glomerular filtration rate, ICI = immune checkpoint inhibitor, ACEi/ARB = angiotensin converting enzyme inhibitor or angiotensin receptor blockade, NSAIDs = nonsteroidal anti-inflammatory drugs, H2B = histone 2 blockade, no. = number, PD1 = programmed death 1, PDL1 = programmed death ligand 1, CTLA4 = cytotoxic T lymphocyte-associated antigen.

| Characteristic | All Patients<br>(N=5004) | Patients surviving ≥1<br>year (N=2563) |
| --- | --- | --- |
| **Demographics** |  |  |
| Age (years) mean (SD) | 64 (13) | 63 (13) |
| Male sex – no. (%) | 2699 (54) | 1377 (54) |
| Race/Ethnicity – no. (%) |  |  |
| White, non-Hispanic | 4529 (91) | 2356 (92) |
| Black, non-Hispanic | 112 (2) | 47 (2) |
| Asian | 157 (3) | 59 (2) |
| Hispanic | 51 (1) | 28 (1) |
| Other or Unknown | 155 (3) | 73 (3) |
| **Coexisting conditions - no. (%)<sup>a</sup>** |  |  |
| Hypertension | 2930 (59) | 1479 (58) |
| Diabetes mellitus | 819 (16) | 427 (17) |
| Cirrhosis | 78 (2) | 36 (1) |
| Chronic obstructive pulmonary disease | 515 (10) | 227 (9) |
| Coronary artery disease | 1168 (23) | 605 (24) |
| Chronic kidney disease |  |  |
| eGFR ≥90 ml/min | 1956 (39) | 959 (37) |
| eGFR 60-89 ml/min | 2249 (45) | 1195 (47) |
| eGFR 45-59 ml/min | 745 (15) | 384 (15) |
| eGFR < 45 ml/min | 52 (1) | 25 (1) |
| **Baseline medications - no. (%)** |  |  |
| ACEi / ARB | 1390 (28) | 670 (26) |
| Uric Acid Lowering agent | 243 (5) | 93 (4) |
| NSAIDs | 2499 (50) | 1256 (49) |
| Diuretics | 1315 (26) | 544 (21) |
| Nephrotoxic antineoplastic therapies | 2081 (42) | 889 (35) |
| Histone 2 receptor blockade | 2616 (52) | 1212 (47) |
| Proton pump inhibitors | 2127 (43) | 929 (36) |
| **ICI Class - no. (%)** |  |  |
| PD1 | 3720 (74) | 1846 (72) |
| PDL1 | 519 (10) | 283 (11) |
| CTLA4 | 355 (7) | 205 (8) |
| Combination CTLA4/PD1 | 410 (8) | 229 (9) |
| **Cancer Type - no. (%)** |  |  |
| Lung | 1706 (34) | 723 (28) |
| Melanoma | 1157 (23) | 810 (32) |
| Head and Neck | 506 (10) | 241 (9) |
| Genitourinary | 365 (7) | 241 (9) |
| Gynecologic | 317 (7) | 153 (6) |
| Gastrointestinal | 243 (5) | 74 (3) |
| Blood* | 196 (4) | 98 (4) |
| Breast | 162 (3) | 64 (3) |
| Sarcoma | 86 (2) | 38 (1) |
| Other | 266 (5) | 119 (5) |

We excluded patients without baseline creatinine, without ≥1 follow-up creatinine, potentially receiving placebo infusion as part of a clinical trial, or on dialysis at ICI start. Death was determined by chart review or was assigned 45 days after the last laboratory datapoint in patients without laboratory studies for >6 months.

### Primary and secondary outcomes

The primary outcome was a composite of new-onset CKD (defined by eGFR < 60 mL/min/1.73m^2^ at least twice separated by 90 days with no intervening values ≥ 60 mL/min/1.73m^2^) or a sustained 30% decline in eGFR compared to baseline for more than 90 days with no interim eGFR reflecting a < 20% decline from the baseline.^18^ A 30% decline in eGFR was chosen based on data indicating that this is predictive of future end-stage renal disease (ESRD).^19^

As a secondary outcome, we defined “rapid eGFR decline” as >3mL/min/1.73m^2^ decline per year, which is associated with ESRD and adverse cardiovascular outcomes.^20, 21^ We determined the average slope of eGFR decline in the one-year prior to ICIs among all patients with a least 12 months of data prior to starting ICIs to determine the change in eGFR slope after beginning ICIs.

### Statistical Analysis

Baseline characteristics were described using means and standard deviations (SD) for continuous variables, and counts and percentages for categorical variables. For the primary outcome we censored the follow-up at the time of composite outcome occurrence, death, or at the end of the follow-up period, whichever happened first. Fine-Gray subdistribution hazard models were performed to identify predictors of the primary outcome accounting for the competing risk of death. We examined both traditional risk factors associated with CKD progression and cancer-related factors (cancer type, ICI class, or prior receipt of nephrotoxic cancer therapies). The final model selection was guided by a combination of clinical plausibility and by information criteria (Akaike and Bayesian information criterion). Covariates in the final model included age, gender, race/ethnicity, baseline eGFR, diabetes, coronary artery disease, and cancer type.

For our secondary outcome, we performed a least squares regression model using all eGFR values available to estimate the slope of eGFR decline for each patient.^21^ The rate of rapid eGFR decline was the proportion of patients whose slope declined >3mL/min/1.73m^2^ per year among all surviving patients. A two-sided P value of <0.05 was considered to be statistically significant. All analyses were performed using SAS 9.4. The Institutional Review Board approved this study and waived the need for informed consent.

## Results

We identified 5934 adult patients who began ICIs at MGB between 2010-2018. After applying the exclusion criteria, 5004 were included, and the 2563 who survived ≥ 1 year were included in the primary analysis (**Figure 1**). Baseline characteristics of the cohort are shown in **Table 1**. Among patients surviving ≥1 year, mean age was 63 years (SD 13), 54% were male, and 92% were white, non-Hispanic. Medical comorbidities were common, including hypertension in 58%, diabetes in 17%, coronary artery disease in 24% and CKD in 16%. The majority (72%) were treated with PD1 inhibitors, followed by 11% with PDL1 inhibitors, 8% with CTLA4 inhibitors and 9% with combination PD1/CTLA4 inhibition (**Figure 2**). The breakdown of cancer type among all participants is show in **Table 1** and broken down by year of ICI administered in **Figure 3**. Patients were followed for a median of 688 days (interquartile range, 496 to 1031).

**Figure 1.**
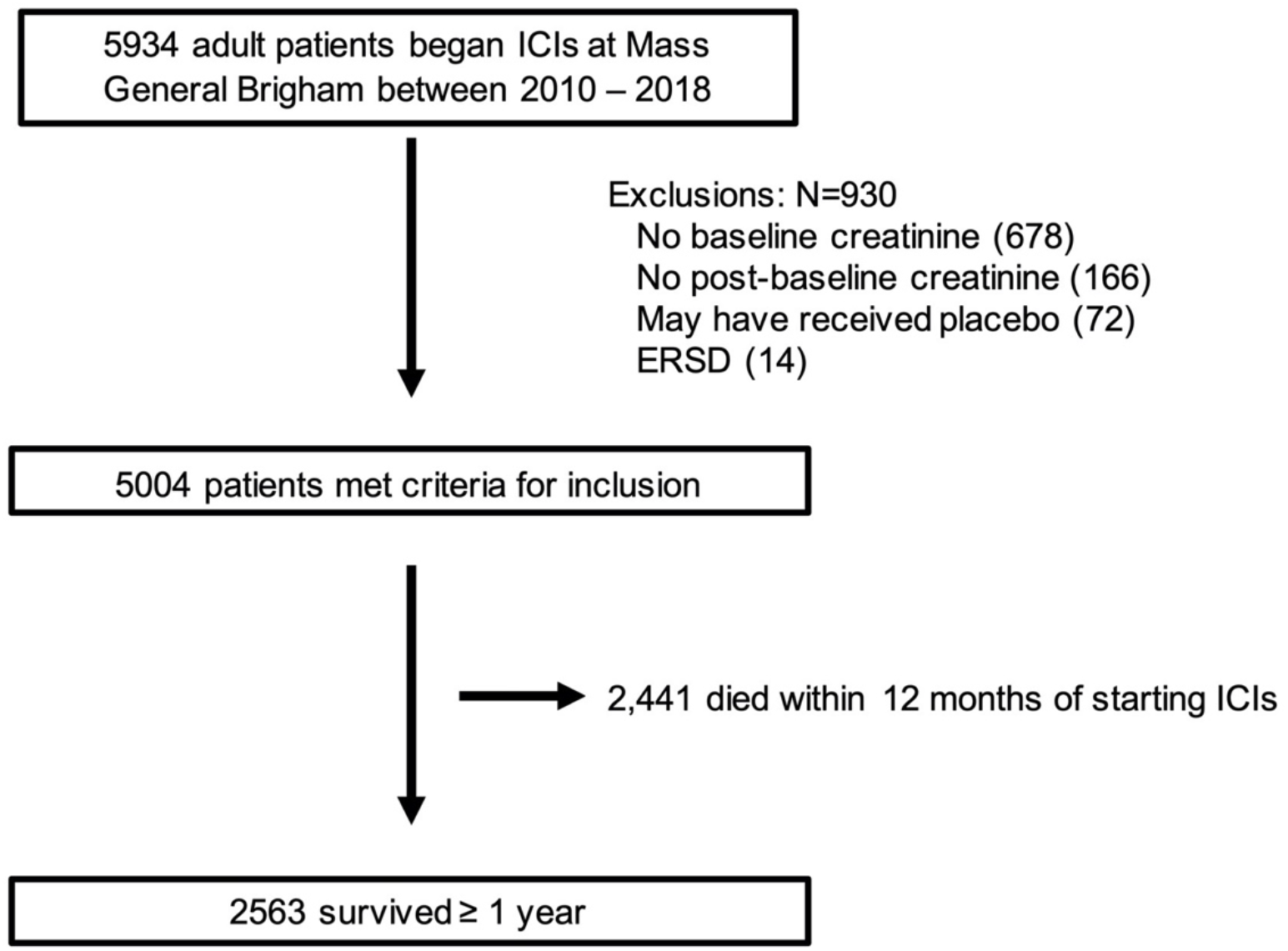
Study cohort. Abbreviations: ESRD = end stage renal disease; ICIs = immune checkpoint inhibitors.

**Figure 2.**
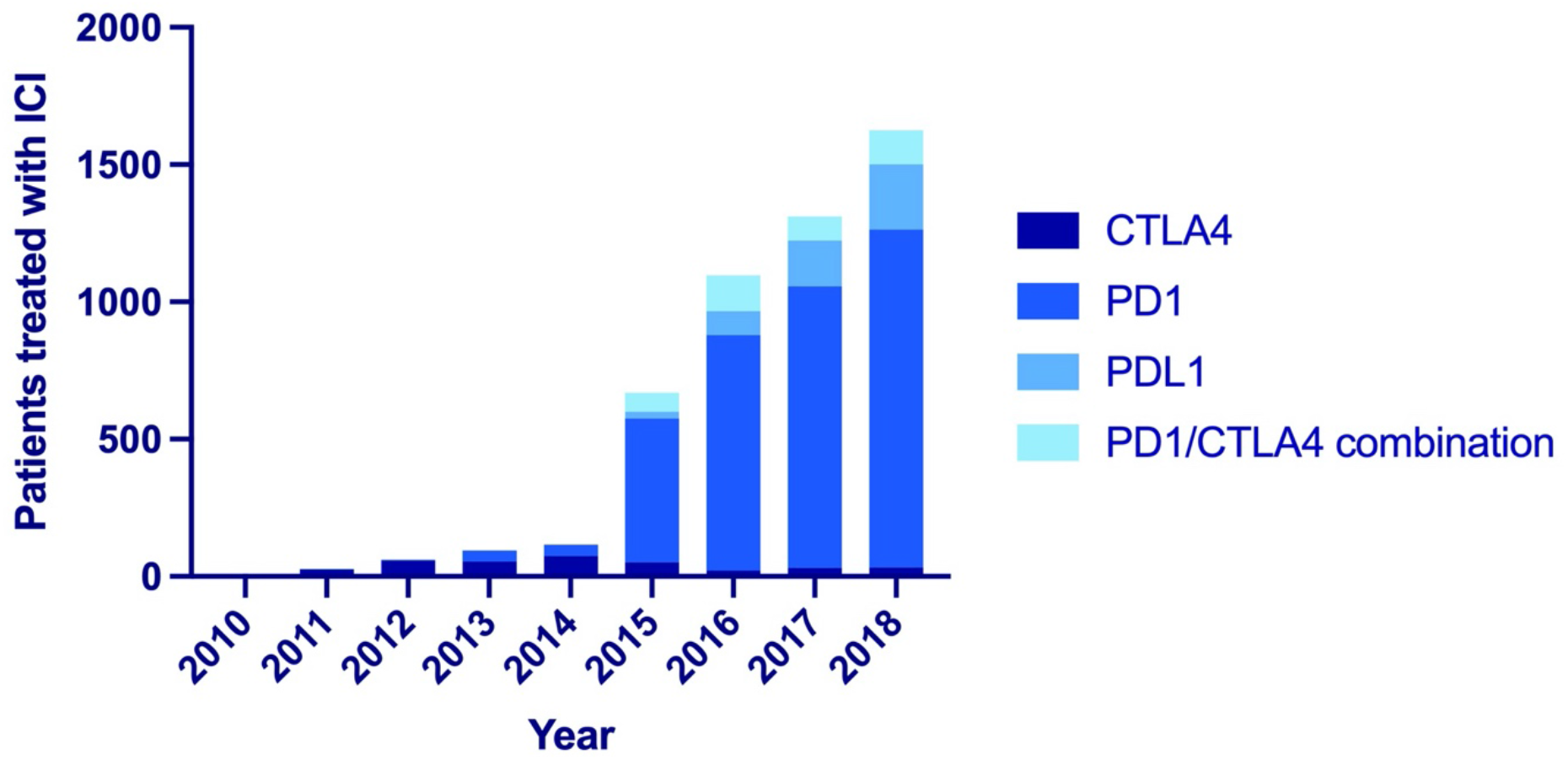
Initial ICI-class used, by year of initiation. Abbreviations: ICI = immune checkpoint inhibitor, CTLA4 = cytotoxic lymphocyte- associated protein 4, PD1 = programmed death 1, PDL1 = programmed death ligand 1

**Figure 3.**
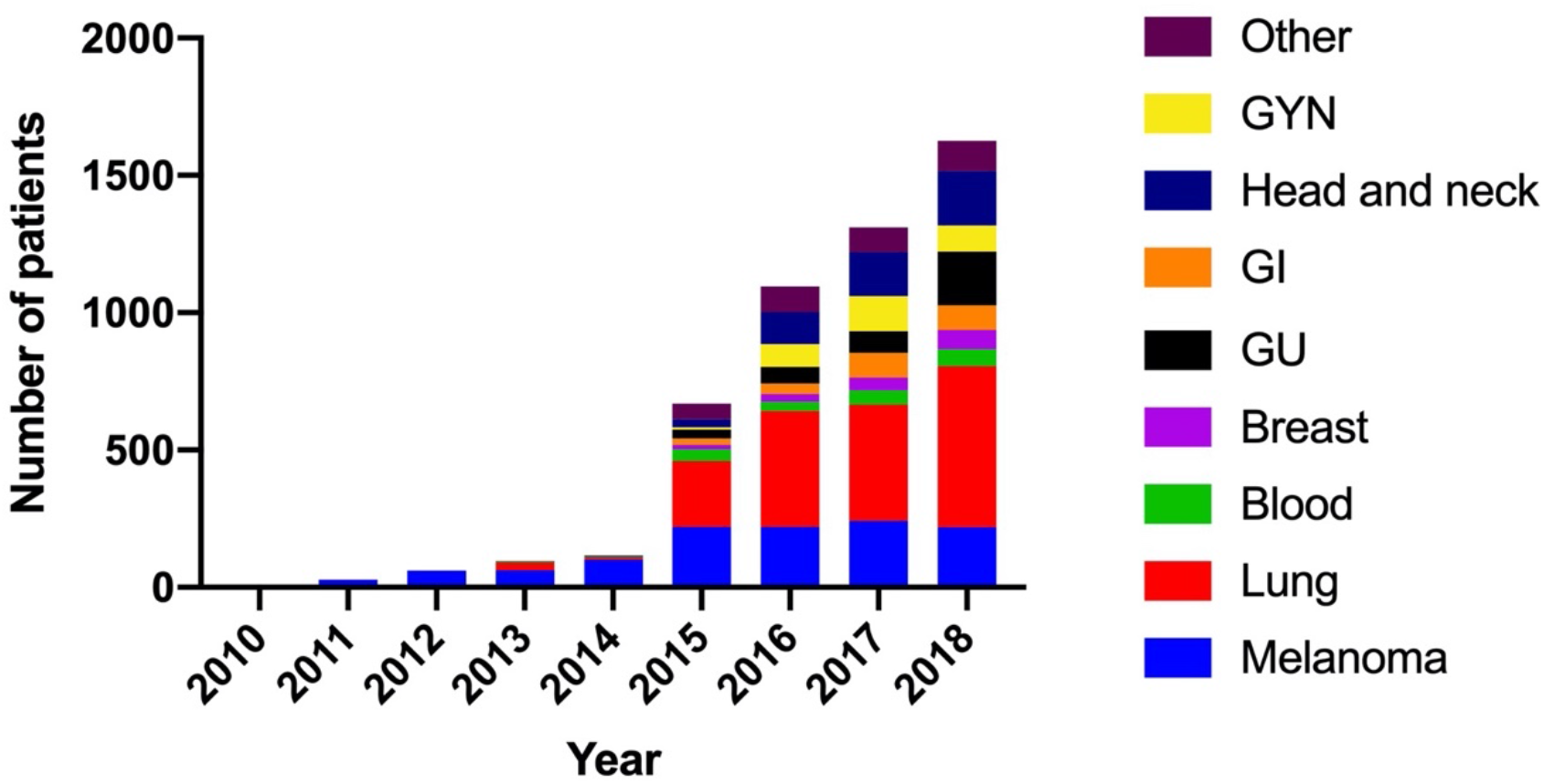
Breakdown of cancer type, by year ICI-initiation. Abbreviations: ICI = immune checkpoint inhibitor; GYN = gynecologic; GI = gastrointestinal; GU = genitourinary.

### Incidence and Predictors of Primary and Secondary Outcomes

The incidence rate of the primary outcome (new-onset CKD or 30% eGFR decline) by each year survived post-ICI is shown in **Figure 4**. Among patients surviving ≥1 year, the primary composite outcome occurred a median of 471 days after the first dose of ICI (IQR, 304-782 days). The adjusted hazard ratios for predictors of the primary outcome are shown in **Figure 5**. Age and coronary artery disease were each independently associated with the primary outcome, adjusted HR (95% CI) 1.12 (1.06, 1.19) per 5 years, and 1.27 (1.00 – 1.62), p < 0.001 and p = 0.049, respectively (**Figure 6**). There were no differences in the incidence of the primary outcome according to ICI class (p = 0.43) (**Figure 7)**.

**Figure 4.**
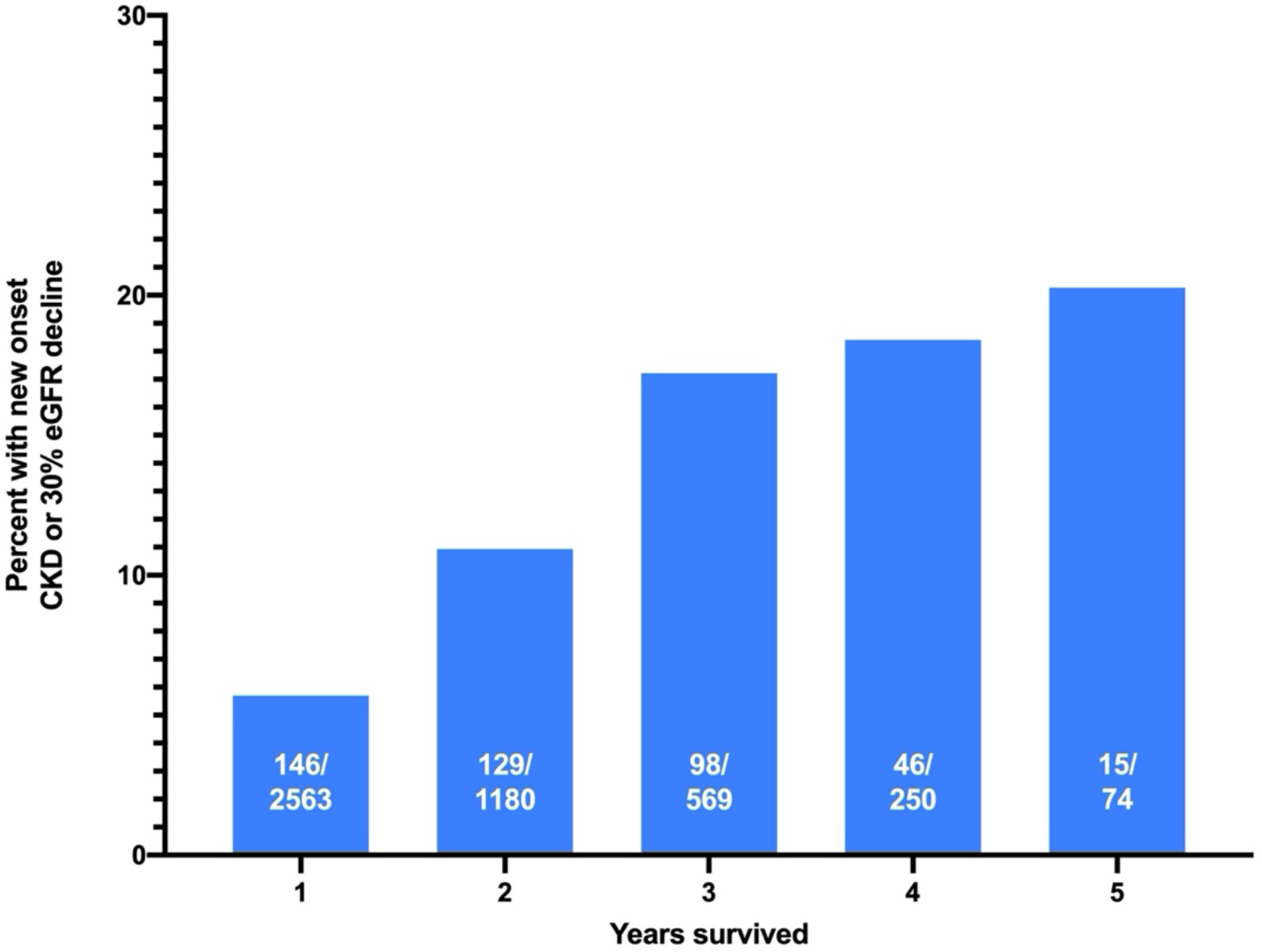
New-onset CKD or sustained 30% eGFR decline among patients surviving ≥ 1yr. Percent experiencing new onset CKD or a sustained 30% eGFR decline among patients surviving 1-5 years. Numerator represents the number of experiencing the primary composite outcome among the total number surviving each year (denominator).

**Figure 5.**
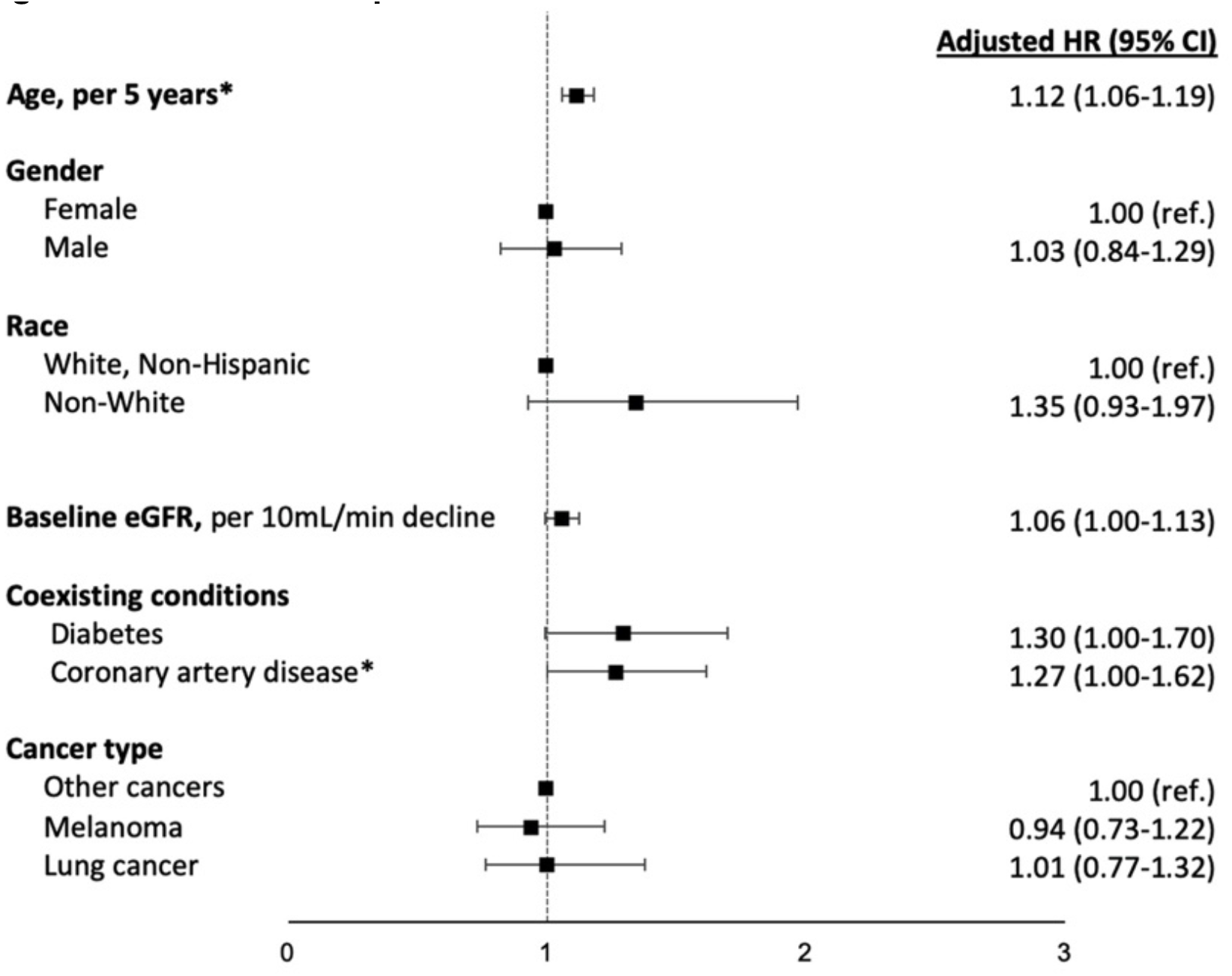
Incidence and predictors of new onset CKD or sustained 30% eGFR decline. Fine and Gray proportional subdistribution hazard models were used to estimate the association between baseline characteristics and the primary composite outcome of new-onset CKD or 30% decline in eGFR among the 2563 patients suriving at least 1 year. Death was determined by chart review or was assigned 45 days after the last laboratory datapoint in patients without laboratory studies for >6 months and was treated as a competing event. The statistically significant variables, age and baseline coronary artery disease, are noted with an asterix. Borderline significant predictors included baseline eGFR and diabetes, p=0.054 and p=0.053 respecitvely. <u>Abbreviations:</u> CI, confidence interval; CKD, chronic kidney disease; HR, hazard ratio; ref., referent.

**Figure 6.**
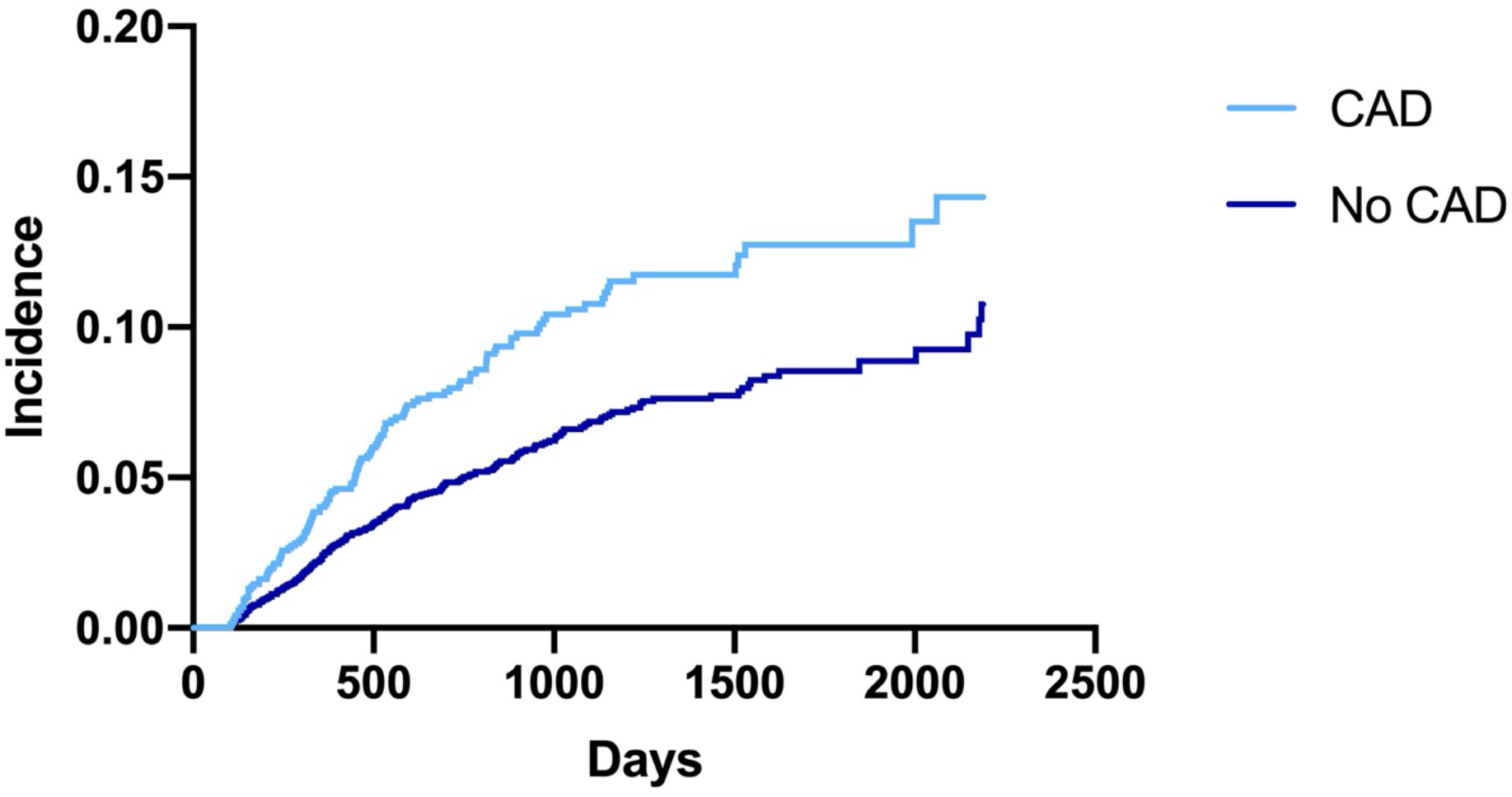
Cumulative incidence of new-onset CKD or sustained 30% eGFR decline among patients with and without baseline coronary artery disease. The cumulative incidence of the primary outcome is shown among the 5004 patients initiating immune checkpoint inhibitors who met criteria for inclusion. At baseline 1168 had coronary artery disease (CAD) and 3,836 did not.

**Figure 7.**
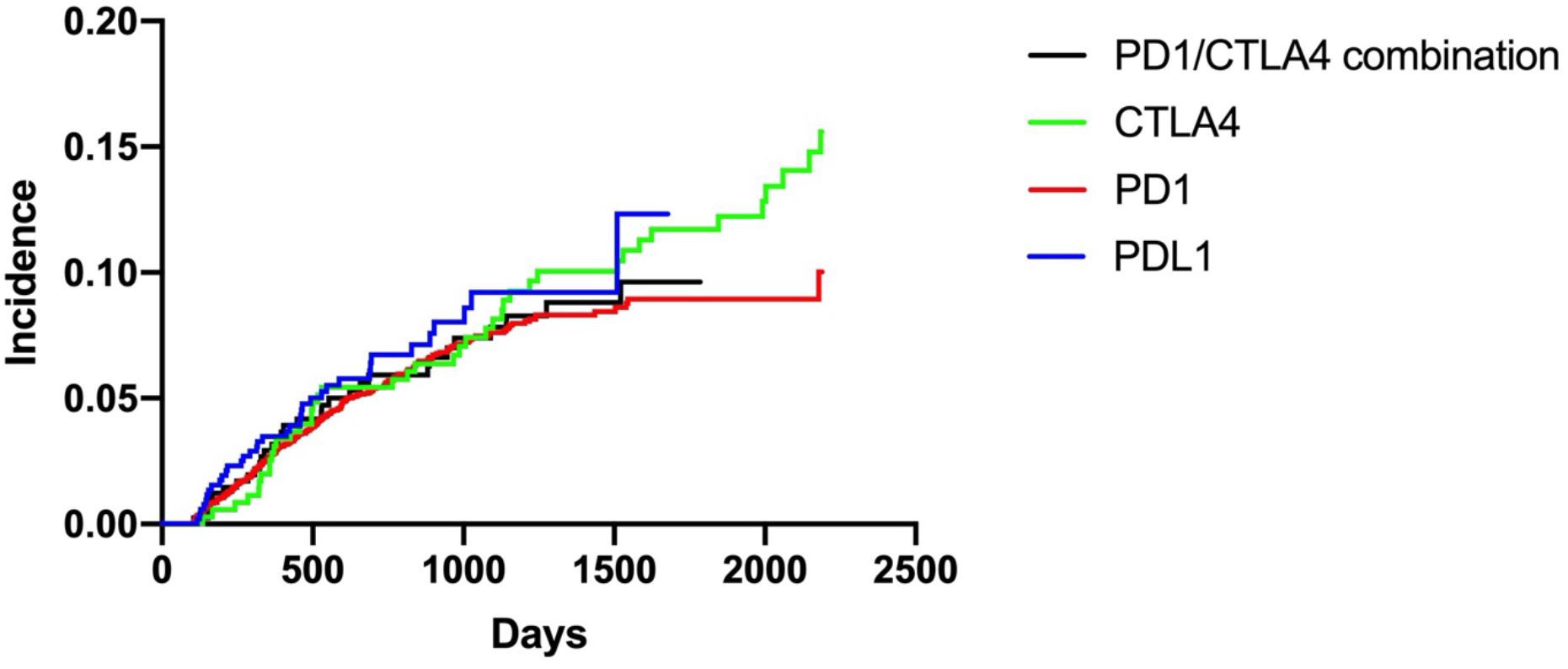
Cumulative incidence of new-onset CKD or sustained 30% eGFR decline among different ICI types. The cumulative incidence of the primary outcome is shown among the 5004 patients initiating immune checkpoint inhibitors who met criteria for inclusion. 3720 patients were treated with PDL1 inhibitors, 519 with PD1 inhibitors, 355 with CTLA4 inhibitors and 410 with combination therapy. Abbreviations: eGFR = estimated glomerular filtration rate; ICI = immune checkpoint inhibitor

The secondary outcome, rapid eGFR decline >3mL/min/1.73m^2^ per year was also common, affecting 35% of those who survived at least 3 years (**Table 2**). Analysis of pre-ICI eGFR slope decline among patients surviving at least one year who had at least 1 year of baseline data (N=1335) showed a significant acceleration of eGFR decline after ICIs (1.4mL/min/year average decline prior to ICI that increased to 3.7mL/min/year decline after ICI, p=0.012) (**Table 3**).

**Table 2.**
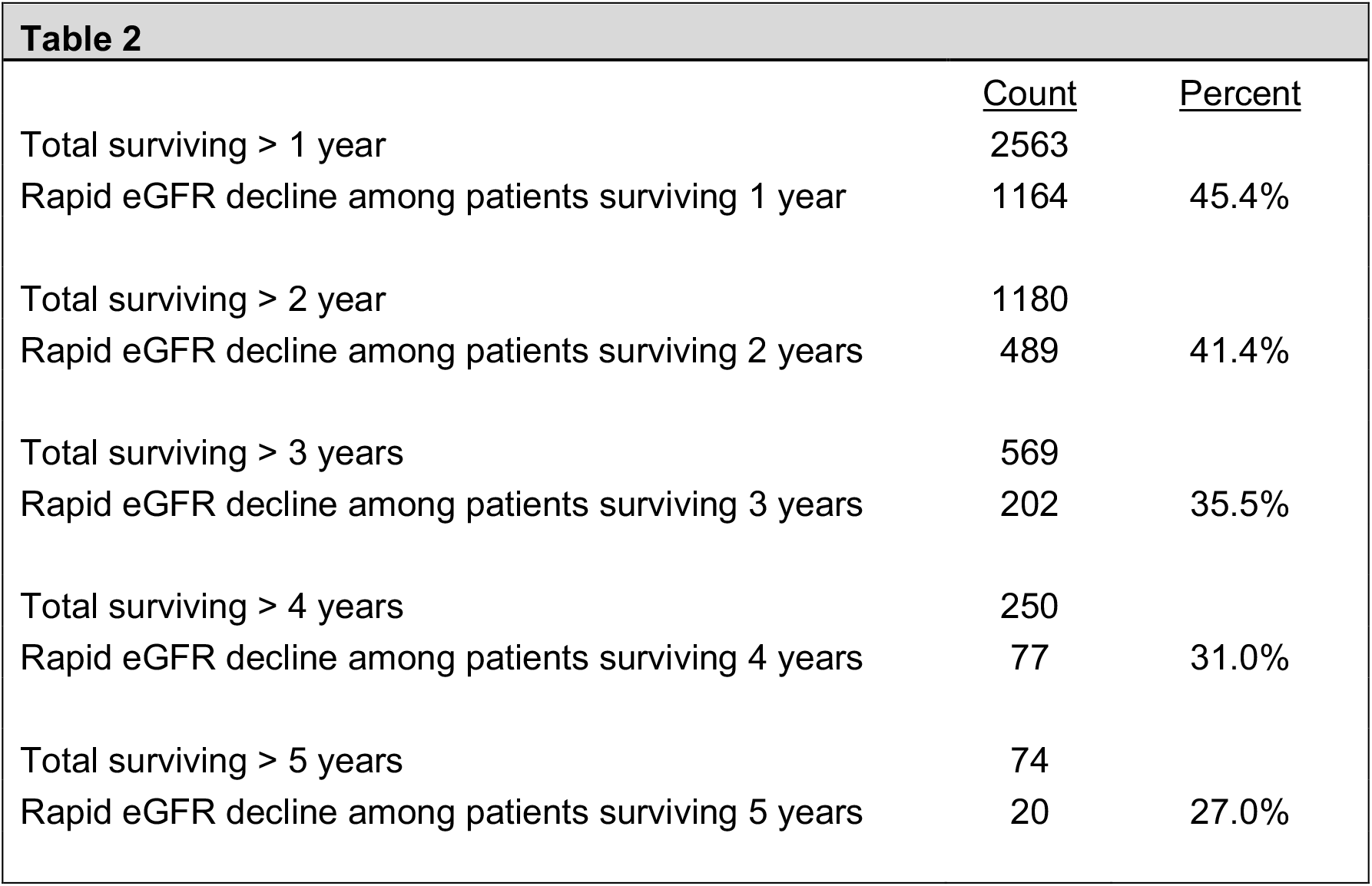
Frequency of rapid eGFR decline (>3mL/min/1.72m^2^ per year decline) in patients surviving ≥1yr.

**Table 3.**
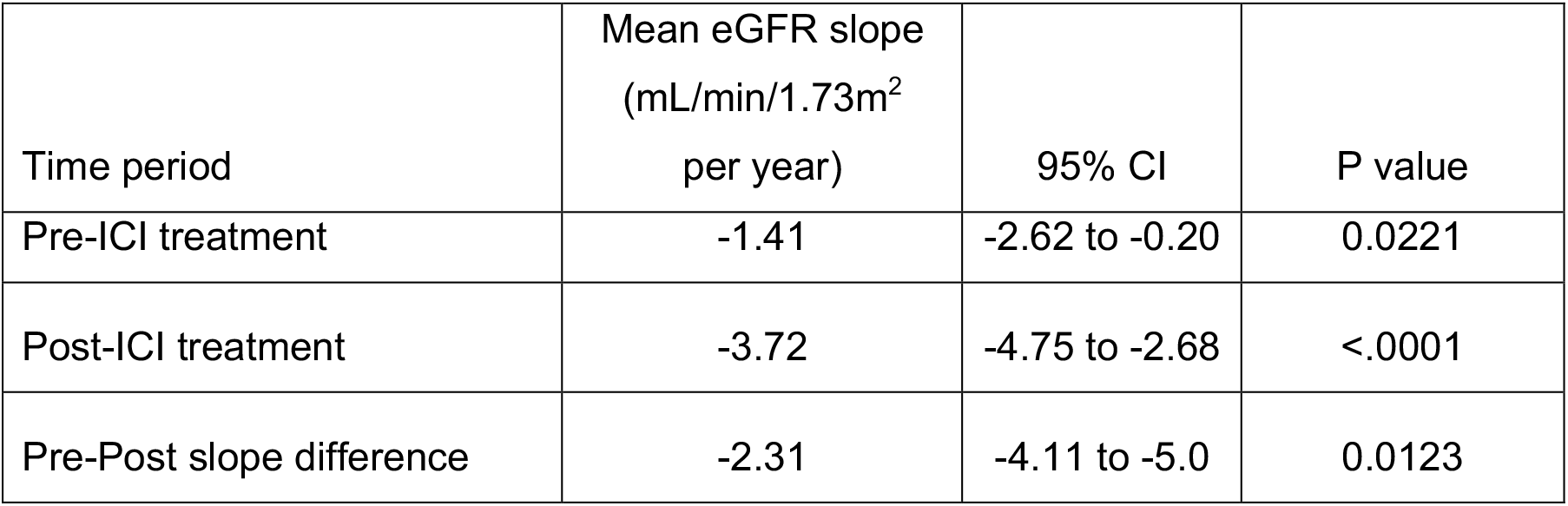
Average eGFR slope one year prior and one year after starting ICIs.

| Time period | Mean eGFR slope<br>(mL/min/1.73m<sup>2</sup><br>per year) | 95% CI | P value |
| --- | --- | --- | --- |
| Pre-ICI treatment | -1.41 | -2.62 to -0.20 | 0.0221 |
| Post-ICI treatment | -3.72 | -4.75 to -2.68 | <.0001 |
| Pre-Post slope difference | -2.31 | -4.11 to -5.0 | 0.0123 |

## Discussion

This retrospective cohort study assessing kidney function in 2362 patients who survived at least one year after treatment with ICIs demonstrates that new-onset CKD or a clinically significant decline in eGFR is common. The robustness of this finding is confirmed by the high prevalence of rapid eGFR decline (>3mL/min/1.73m^2^ per year) and by analysis of eGFR slope decline before and after initiating ICIs.

Our study is descriptive and is limited by the absence of a control group. However, given the extremely limited survival of many metastatic cancers, such as melanoma, prior to ICIs, it is not possible to find an adequately matched population with metastatic cancers with the potential for long-term survival. When patients served as their own control by comparing pre and post- treatment eGFR slope, a substantial acceleration of eGFR decline was noted. Prior large studies of kidney function in patients with cancer evaluated eGFR cross-sectionally among patients with undergoing chemotherapy and found approximately 50-60% have renal impairment (eGFR < 90 mL/min/1.73m^2^) or CKD.^22-24^ We found similar prevalences at baseline, but noted incidence CKD in substantial numbers. Since patients with cancer lose muscle mass over time, the true burden of CKD may be underestimated. Our study needs to be validated in this and other cohorts with longer follow-up.

The high prevalence of new onset CKD and eGFR decline that occurs after ICIs may have crucial health consequences for cancer survivors and may limit the use or dosage of other anticancer drugs and eligibily for clinical trial enrollment.^25, 26^ Future studies will be needed to determine the pathophysiology of CKD after ICIs and evaluate strategies to slow eGFR decline.

Our findings shed light on the long-term kidney disease burden among the growing population of cancer survivors exposed to ICIs.

## Data Availability

De-identified data can be made available upon request.

---

**Table 4.** Among the 2363 patients who survived > 1 year, a total of 1335 had been followed with serial eGFR measurements for at least 1 year prior to initiating ICIs. Pre-post slope difference is compared with a paired T-test

