## Supplemental Table 1 for "Clinically significant estimated glomerular filtration rate decline is common in patients receiving immune checkpoint inhibitors: implications for long-term cancer survivors"

**Supplemental Table 1. List of nephrotoxic anti-cancer therapies**

Nephrotoxic chemotherapies

cisplatin

oxaliplatin

carboplatin

gemcitabine

methotrexate

ifosfamide

lomustine

streptozocin

carmustine

clofarabine

pemetrexed

doxorubicin

arsenic

Nephrotoxic targeted agents

Crizotinib

alectinib

lorlatinib

dasatinib

bosutinib

imatinib

Trametinib

Cobimetinib

Cetuximab

afatinib

erlotinib

gefitinib

Trastuzumab

pertuzumab

temsirolimus

sirolimus

lenvatinib

regorafenib

vandetanib

ibrutinib

olaparib

selinexor

vemurafenib

bortezomib

encorafenib

ifosfamide

dabrafenib

carfilzomib

ixazomib

axitinib

cabozantinib

pazopanib

sorafenib

sunitinib

Bevacizumab

venetoclax

Nephrotoxic immunomodulators

lenalidomide

pomalidomide

thalidomide
